# High Burden of Ventricular Arrhythmias and Cardiac Conduction Disease in Pediatric Patients with Pathogenic Desmin (*DES*) Variants

**DOI:** 10.1101/2025.09.29.25336933

**Authors:** Babken Asatryan, Marina Rieder, Brittney Murray, Steven A. Muller, Crystal Tichnell, Alessio Gasperetti, Richard T. Carrick, Emily Joseph, Doris G. Leung, Anneline S.J.M. te Riele, Stefan L. Zimmerman, Hugh Calkins, Cynthia A. James, Caridad M. De La Uz, Andreas S. Barth

## Abstract

**Background:** Pathogenic variants in the desmin gene (*DES*) are implicated in diverse cardiomyopathy and skeletal myopathy phentoypes with high rates of major adverse cardiac events (MACE). Although pediatric cases have been reported, the genetic profile, clinical features and outcomes in affected children remain poorly defined. This study aimed to comprehensively characterize these aspects in the pediatric population.

**Methods:** We conducted a systematic review and individual patient data meta-analysis of Medline (PubMed) and Embase, including patients with pathogenic or likely pathogenic *DES* variant(s) diagnosed before age 21. MACE were defined as cardiac conduction disease (CCD) requiring device implantation, sustained ventricular arrhythmias (VA), and heart failure (HF) events.

**Results:** Fifty-one pediatric patients were included (58.8% male; 64.7% probands; median age at first evaluation: 14.0 years [IQR 11.0–17.0], median follow-up: 5.0 years [1.0–11.5]). Among probands, 60.6% (20/33) had a heterozygous variant (20 non-frameshift/non-truncating and 2 frameshift/truncating), while the remaining 11 (33.3%) harbored biallelic genotypes (homozygous (n=8), compound heterozygous (n=3)). At presentation, 54.9% (28/51) patients had cardiomyopathy and 54.9% had skeletal myopathy. CCD occurred in 39.2% (median age: 19.0 years), sustained VA in 21.6% (median age: 18.0 years), and HF events in 37.3% (median age: 19.0 years). In total, 64.7% experienced MACE (median age: 17.0 years [13.0–19.0]), and 19.6% died during follow-up (median age: 23.5 years [14.8–28.0]). Rates of cardiomyopathy diagnosis, CCD, VA and HF events as well as composite MACE were similar between those with a heterozygous and biallelic variants (p>0.05 for all), but the latter group had nearly twice higher rate of skeletal myopathy (43.6% vs 83.3%, p=0.016).

**Conclusion:** Pediatric *DES-*associated disease is characterized by heterogeneous cardiomyopathy phenotypes and substantially high MACE burden. Probands were more likely than genotype-positive relatives to develop CCD and HF events, while sustained VA rates were similar and substantial in both groups. Cardiomyopathy rates and outcomes were comparable between patients with heterozygous and biallelic variants.

**WHAT IS KNOWN?:**

- Desmin (*DES*)-associated cardiomyopathy is a rare genetic disorder characterized by variable cardiac and skeletal muscle involvement, typically presenting in adulthood with conduction system disease, ventricular arrhythmias, or heart failure.
- While case reports suggest that pediatric onset is possible, the clinical features, genetic landscape, and outcomes in children remain poorly characterized.

**WHAT THE STUDY ADDS:**

- In pediatric patients with *DES* variants, cardiac phenotype expression can commence in early childhood and rapidly progresses with age, with two-thirds of patients experiencing major adverse cardiac events by young adulthood, highlighting the critical need for initiation of comprehensive cardiac evaluation in early childhood in patients with *DES* variants.
- Cardiac conduction disease and heart failure were the most common events in pediatric *DES* patients, especially in probands, though arrhythmic risk was present, and substantial, both in probands and relatives.
- Multisystem involvement, including skeletal muscle symptoms, was common in pediatric *DES* patients, particularly those with biallelic variants, underscoring the importance of early genetic testing and longitudinal, multidisciplinary surveillance in at-risk children, regardless of family history.

## INTRODUCTION

Desminopathies encompass a spectrum of cardiomyopathy and skeletal myopathy phenotypes caused by pathogenic and likely pathogenic variants in the desmin (*DES*) gene, which may manifest either in isolation or combination.^1,2^ Desmin is a muscle-specific intermediate filament protein that forms a cytoskeletal network linking the contractile apparatus to other structural elements of the cell, thus providing maintenance of the cellular integrity, enabling force transmission, and facilitating mechanochemical signaling.^3^ Cardiac involvement in *DES-*related disease is notably variable and associated with a high burden of major adverse cardiac events (MACE), such as cardiac conduction disease (CCD) requiring pacemaker implantation, sustained ventricular arrhythmias (VA), and heart failure (HF) events.^4^ Although symptoms typically emerge in the third or fourth decade of life,^2,4^ disease onset and severity vary widely, and pediatric cases with diverse manifestations have been reported.^5–8^ Pediatric cardiomyopathies are generally associated with poorer outcomes than their adult-onset counterparts, with earlier diagnosis linked to worse prognosis.^9,10^ However, the rarity and clinical heterogeneity of DES-related disease, which frequently leads to presentation across multiple specialties, have limited systematic characterization in children.^11^

This study sought to comprehensively characterize the genetic landscape, phenotypic spectrum and clinical outcomes of pediatric patients with *DES* variants, leveraging the pediatric subset of a recent systematic review and individual patient data meta-analysis of patients with pathogenic *DES* variants.^4^ This effort aligns with growing recognition of the importance of early genetic diagnosis in cardiomyopathies, which can inform surveillance strategies, guide therapeutic interventions, and facilitate family counseling.^12^

## METHODS

### Compliance with Ethical Standards

The data used in the systematic review and individual patient data meta-analysis are publicly available and institutional review board or ethics committee approval was not required. The dataset generated through the extraction of data from published studies will be made available upon reasonable request.

### Data Source and Search Strategy

We conducted a systematic review and individual patient data meta-analysis of Medline (PubMed) and Embase, including studies reporting at least one patient with a pathogenic or likely pathogenic *DES* variant. Detailed methods, including data source, search strategy, study selection, data extraction, and data synthesis, have been reported previously.^4^ This study focused on the pediatric subset of that cohort.

### Patient population—inclusion and exclusion criteria

Patients were included if they:

i. harbored a pathogenic and likely pathogenic *DES* variant, as determined at core lab adjudication;^13^
ii. were alive and <21 years of age at the time of the first evaluation; and
iii. had at least one clinical encounter with cardiac evaluation (at least ECG and either echocardiogram or cardiac magnetic resonance imaging.

Patients were excluded from the study if they met any of the following criteria:

i. had a confirmed or probable digenic inheritance (i.e. carried additional variant(s) of uncertain significance, pathogenic and likely pathogenic variant in another gene implicated in inherited arrhythmias or cardiomyopathies); or
ii. experienced a major adverse cardiac event (MACE) due to a presumed alternative cause (e.g. ventricular fibrillation in the context of acute myocardial infarction).

Demographic, clinical, genetic data, and family history information were abstracted. Diagnosis of cardiomyopathy relied on applicable clinical practice guidelines and recommendations; ascertainments of distinct cardiomyopathy phenotypes by original reports were accepted as meeting the corresponding diagnostic criteria. Sustained VA was defined as a composite of the occurrence of sustained ventricular tachycardia, appropriate ICD interventions, ventricular fibrillation/flutter, and sudden cardiac arrest / sudden cardiac death (SCD) episodes, as reported. CCD events were defined as second-degree Mobitz type II, advanced degree or complete AV block, any symptomatic AV block or other CCD that led to CIED implantation. HF events were defined as a composite of hospitalization for HF, left ventricular assist device (LVAD) implantation/cardiac transplant, and HF-related death. Rates of CCD requiring cardiac implantable electronic device (CIED) placement, sustained VA, HF events, and composite MACE were assessed.

### Statistical Analysis

Continuous, normally distributed variables were summarized using mean (SD) and compared using 2-sample Student’s t-tests. Non-normally distributed continuous data were summarized using median and interquartile range and compared using Wilcoxon-rank sum tests. Categorical data were summarized using percentages and compared using χ^2^ or Fisher exact tests, as appropriate. α was set at 0.05 for statistical significance. Data were analyzed using R statistical computing (version 4.5.1).^14^

## RESULTS

### Demographic and Clinical Characteristics

A total of 51 pediatric patients with pathogenic or likely pathogenic *DES* variants were included, comprising 22.2% of our entire cohort in our systematic review and individual patient data meta-analysis of *DES* patients (**Figure 1**). Of these, 30 (58.8%) were male and 33 (64.7%) were probands (**Table 1**). Compared with adults, pediatric patients were more likely to be the proband in the family (64.7% (33/51) vs 48.6% (87/179), p=0.042). The median age at first evaluation was 14.0 years (IQR 11.0–17.0), with a median follow-up duration of 5.0 years (IQR 1.0–11.5). A family history of SCD (n=7) or sustained ventricular arrhythmia (n=2) was reported in 9 probands (17.6%), and 12 probands (23.5%) had a family history of cardiomyopathy. At initial presentation, the predominant manifestation was cardiac in 26 patients (51.0%), neuromuscular in 13 (25.5%), and both cardiac and neuromuscular in 3 (5.9%); 7 patients (13.7%) were asymptomatic with no cardiac or neuromuscular symptoms.

**Figure 1.**
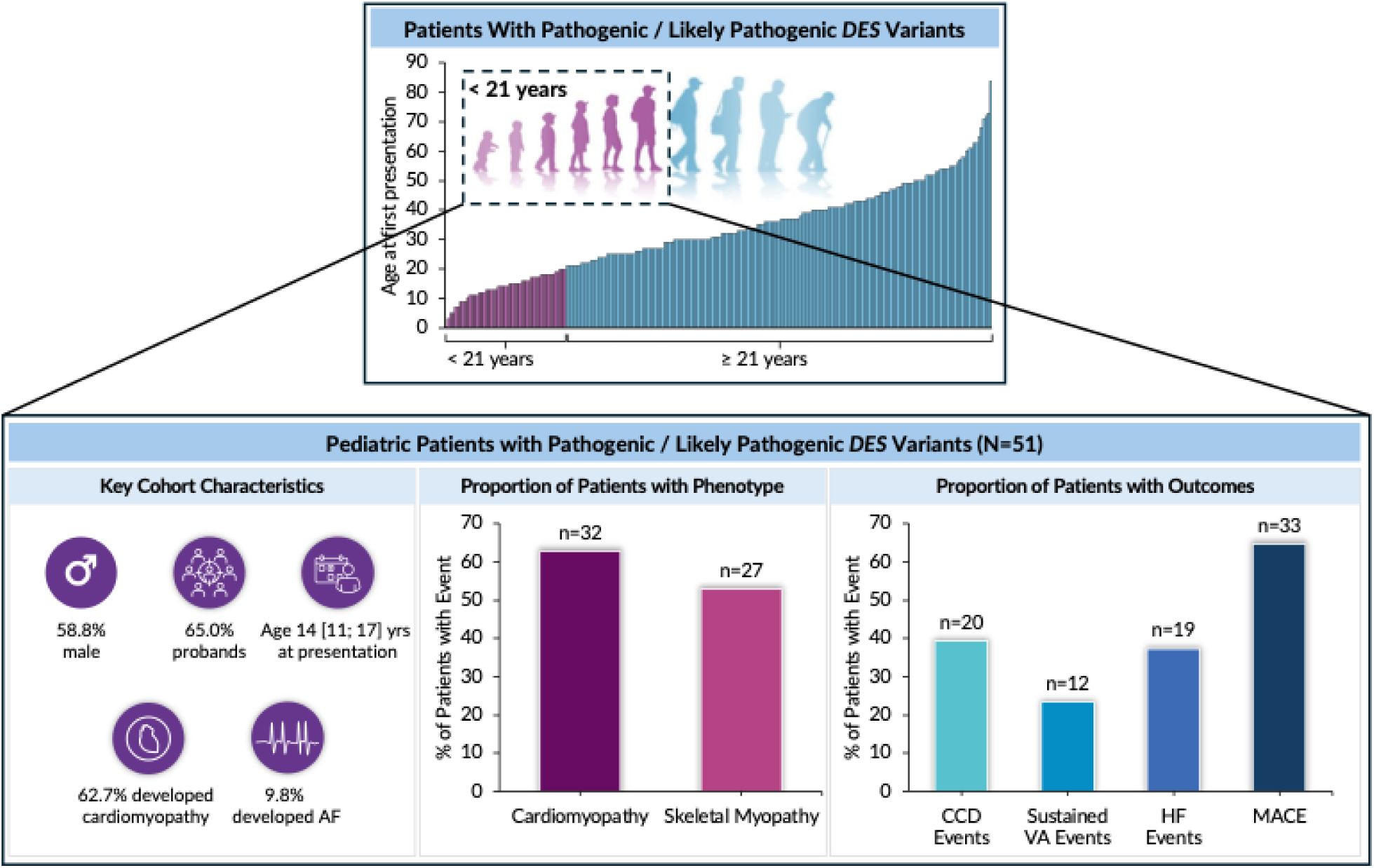
(Graphical Abstract).

**Table 1.**
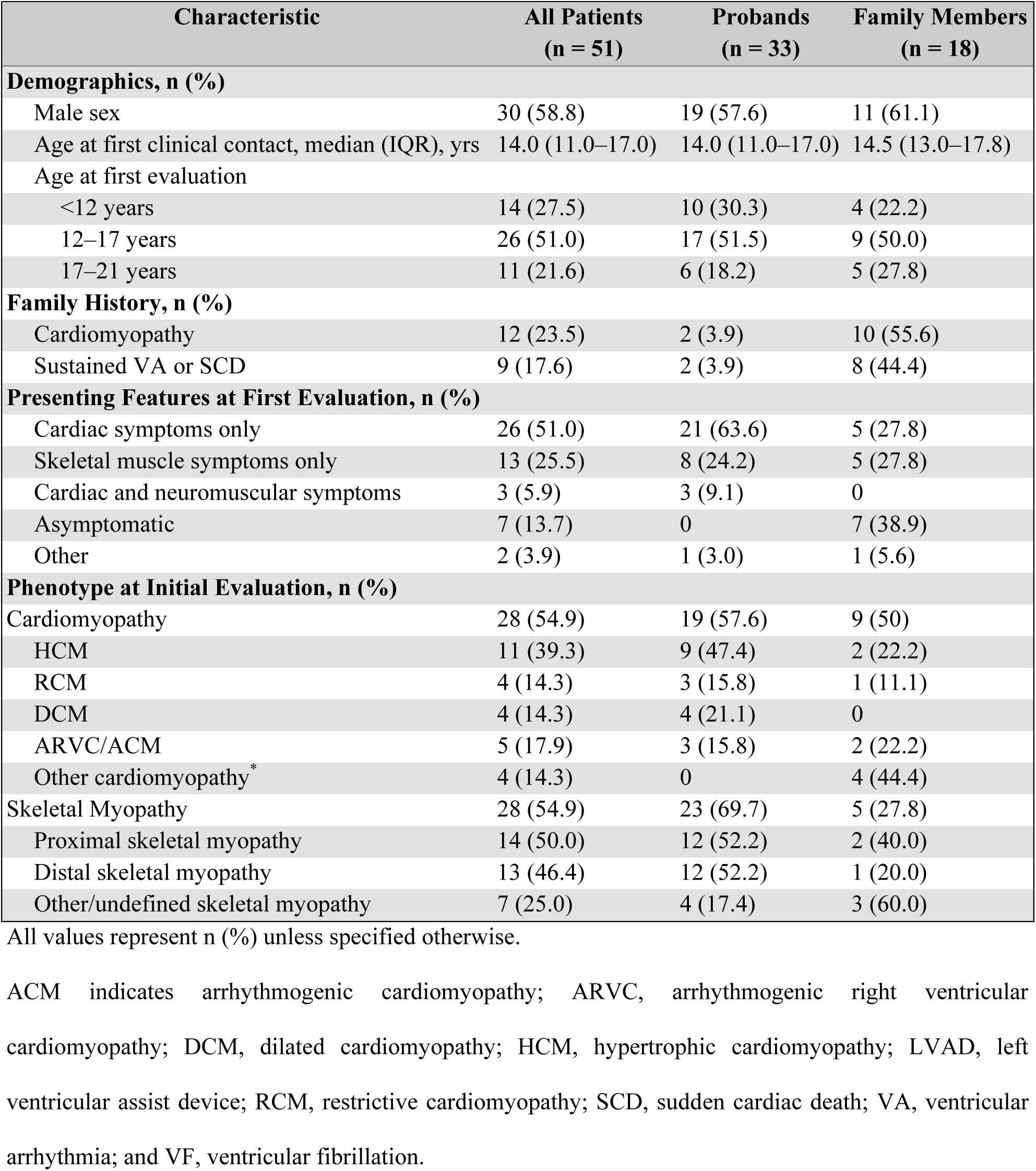
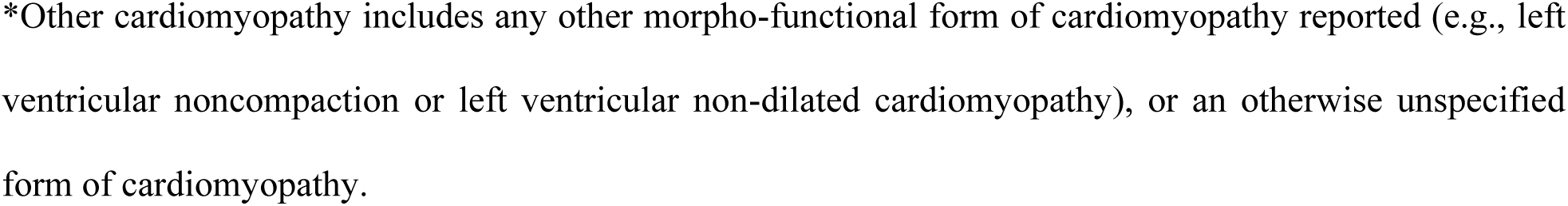
Baseline Demographics and Clinical Characteristics of Pediatric Patients With *DES* Variants.

| Characteristic | All Patients<br>(n = 51) | Probands<br>(n = 33) | Family Members<br>(n = 18) |
| --- | --- | --- | --- |
| **Demographics, n (%)** |  |  |  |
| Male sex | 30 (58.8) | 19 (57.6) | 11 (61.1) |
| Age at first clinical contact, median (IQR), yrs | 14.0 (11.0–17.0) | 14.0 (11.0–17.0) | 14.5 (13.0–17.8) |
| Age at first evaluation |  |  |  |
| <12 years | 14 (27.5) | 10 (30.3) | 4 (22.2) |
| 12–17 years | 26 (51.0) | 17 (51.5) | 9 (50.0) |
| 17–21 years | 11 (21.6) | 6 (18.2) | 5 (27.8) |
| **Family History, n (%)** |  |  |  |
| Cardiomyopathy | 12 (23.5) | 2 (3.9) | 10 (55.6) |
| Sustained VA or SCD | 9 (17.6) | 2 (3.9) | 8 (44.4) |
| **Presenting Features at First Evaluation, n (%)** |  |  |  |
| Cardiac symptoms only | 26 (51.0) | 21 (63.6) | 5 (27.8) |
| Skeletal muscle symptoms only | 13 (25.5) | 8 (24.2) | 5 (27.8) |
| Cardiac and neuromuscular symptoms | 3 (5.9) | 3 (9.1) | 0 |
| Asymptomatic | 7 (13.7) | 0 | 7 (38.9) |
| Other | 2 (3.9) | 1 (3.0) | 1 (5.6) |
| **Phenotype at Initial Evaluation, n (%)** |  |  |  |
| Cardiomyopathy | 28 (54.9) | 19 (57.6) | 9 (50) |
| HCM | 11 (39.3) | 9 (47.4) | 2 (22.2) |
| RCM | 4 (14.3) | 3 (15.8) | 1 (11.1) |
| DCM | 4 (14.3) | 4 (21.1) | 0 |
| ARVC/ACM | 5 (17.9) | 3 (15.8) | 2 (22.2) |
| Other cardiomyopathy* | 4 (14.3) | 0 | 4 (44.4) |
| Skeletal Myopathy | 28 (54.9) | 23 (69.7) | 5 (27.8) |
| Proximal skeletal myopathy | 14 (50.0) | 12 (52.2) | 2 (40.0) |
| Distal skeletal myopathy | 13 (46.4) | 12 (52.2) | 1 (20.0) |
| Other/undefined skeletal myopathy | 7 (25.0) | 4 (17.4) | 3 (60.0) |

All values represent n (%) unless specified otherwise.
ACM indicates arrhythmogenic cardiomyopathy; ARVC, arrhythmogenic right ventricular cardiomyopathy; DCM, dilated cardiomyopathy; HCM, hypertrophic cardiomyopathy; LVAD, left ventricular assist device; RCM, restrictive cardiomyopathy; SCD, sudden cardiac death; VA, ventricular arrhythmia; and VF, ventricular fibrillation.
\*Other cardiomyopathy includes any other morpho-functional form of cardiomyopathy reported (e.g., left ventricular noncompaction or left ventricular non-dilated cardiomyopathy), or an otherwise unspecified form of cardiomyopathy.

Among probands, 10 presented before age 12, 17 during adolescence (12–17 years), and 6 in young adulthood (18–21 years), with cardiomyopathy evident at first evaluation in 4 (40%), 10 (59%), and 5 (83%) individuals, respectively. Among family members, 4 were evaluated before age 12, 9 during adolescence, and 5 in young adulthood, with cardiomyopathy present at first evaluation in 1 (25%), 7 (78%), and 1 (20%) individual, respectively. Before the age of 12, none of the patients in our study experienced sustained VA, 4 patients were diagnosed with CCD at ages 8.5, 9.9, 11, and 11.5 years, while two patients experienced HF events at 6 months and 11 years of age. The youngest relative identified with cardiomyopathy through cascade screening and subsequent clinical evaluation was 7 years old.

By the end of follow-up, 62.7% (32/51) patients had developed cardiomyopathy. Atrial fibrillation (AF) was diagnosed in 5 patients during follow-up, with a median age at first AF diagnosis of 23.0 years (IQR 14.0–24.0).

### Genetic Landscape

Overall, 39 out of 51 patients (76.5%) were identified with a heterozygous variant (including 3 frameshift/truncating variants), while 12 (23.5%) harbored biallelic variants, either compound heterozygous (n=3) or homozygous (n=9). Among probands, 20 of 33 (60.6%) had a heterozygous non-frameshift/non-truncating variant, 2 (6.1%) had a heterozygous frameshift/truncating variant, and 11 (33.3%) biallelic variants with (8 homozygous, 3 compound heterozygous) (**Figure 2**). Biallelic genotypes were significantly more frequent in probands compared to genotype-positive relatives (33.3% (11/33) vs 5.6% (1/18), p=0.025).

**Figure 2.**
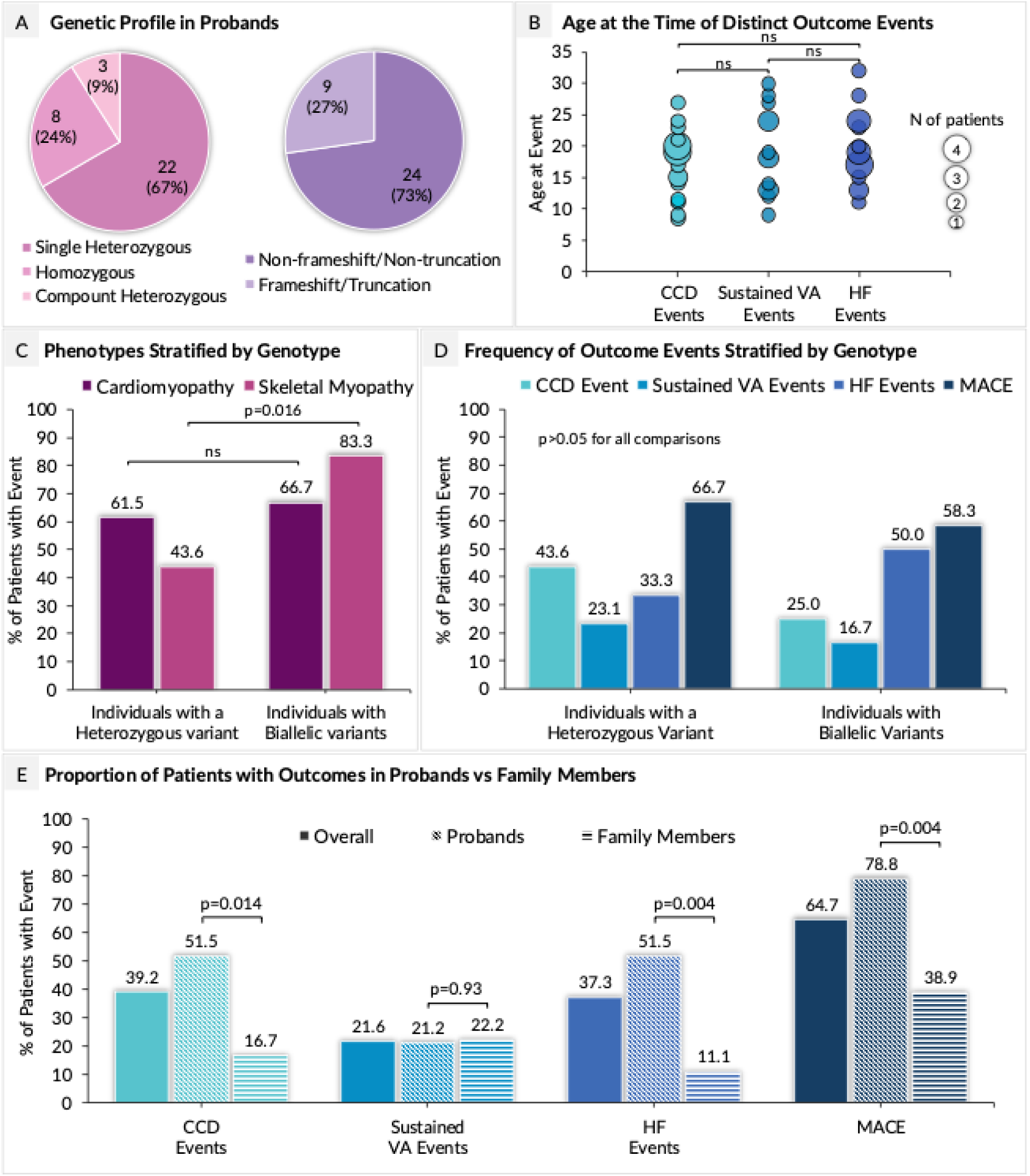
Genetic Profile, Phenotypic Expression, and Major Adverse Cardiac Event Burden in Pediatric Patients with *DES*-Associated Disease. **(A)** Genetic profiles of probands showing a predominance of heterozygous genotypes, and non-frameshift/non-truncating variant class. (**B**) Age at onset was similar across conduction disease (CCD), sustained ventricular arrhythmias (VA), and heart failure (HF); no significant differences observed. (**C**) Cardiomyopathy penetrance was similar in both genotype groups, whereas skeletal myopathy was nearly twice as common in those with biallelic genotypes. (**D**) Frequencies of CCD, VA, HF, and major adverse cardiac events (MACE) were comparable between genotype groups. (**E**) Compared to family members, probands had a significantly higher burden of CCD, HF, and composite MACE, whereas rates of sustained VA events were similar. Color coding for event types in panel E corresponds to the legend shown in panel D.

### Cardiomyopathy and Skeletal Myopathy Phenotypes

Among the 32 patients with cardiomyopathy, the most common phenotypes were dilated (DCM, n=10), restrictive (RCM, n=7), and hypertrophic cardiomyopathy (HCM, n=7). Less frequent were arrhythmogenic cardiomyopathy (ACM, n=4) and arrhythmogenic right ventricular cardiomyopathy (ARVC, n=1), while in 3 patients the cardiomyopathy subtype was not clearly defined. Cardiomyopathy penetrance was similar between patients with heterozygous variant vs those with biallelic genotypes (61.5% and 66.7%, respectively, p=0.748), whereas skeletal myopathy rates were higher in those with biallelic variants (43.6% vs 83.3%, p=0.016).

### Major Adverse Cardiac Events (MACE): CCD, Sustained VA and HF Events

Cardiac events were common in the pediatric *DES* population. CCD requiring CIED placement occurred in 20 of 51 patients (39.2%), with a median age of 19.0 years (IQR 14.8–20.0) at CIED implantation. Sustained VA were observed in 11 patients (21.6%), with a median age at first event of 18.0 years (IQR 13.5–25.5). HF events occurred in 19 patients (37.3%), with a median age at onset of 19.0 years (IQR 16.0–23.5). Overall, 33 patients (64.7%) experienced at least one MACE, with a median age at first MACE of 17.0 years (IQR 13.0–19.0). During follow-up, 10 patients (19.6%) died at a median age of 23.5 years (IQR 14.8–28.0). Causes of death included HF (n=5), SCD (n=2), and respiratory failure from progressive muscle weakness (n=1); the cause was unknown in 2 cases.

Neither distinct outcomes, nor composite MACE rates were statistically different between patients with a heterozygous variant and those with biallelic genotypes (homozygous or compound heterozygous) (p >0.05 for all comparisons) (**Figure 2**).

## DISCUSSION

### Key Findings

Pediatric cardiomyopathies represent a clinically and genetically heterogeneous group of disorders that often follow a more aggressive course than their adult-onset counterparts. Despite the growing number of genetically evaluated pediatric cardiomyopathy cohorts, the clinical phenotypes and outcomes associated with specific genetic forms of pediatric cardiomyopathies remain poorly characterized. In this systematic review and individual patient data meta-analysis, we provide the most comprehensive characterization to date of pediatric *DES*-associated cardiomyopathy. Our findings show that *DES*-related disease can manifest in early childhood, often with cardiac or neuromuscular symptoms, and is associated with a high burden of MACE by young adulthood. Over 60% of affected individuals developed cardiomyopathy, and two-thirds experienced MACE during follow-up. The age distribution at presentation and first phenotypic expression, which is prepubertal in some and delayed into adolescence or early adulthood in others, underscores the variable penetrance and progressive nature of *DES* cardiomyopathy. Notably, biallelic variants (compound heterozygous or homozygous) were common in pediatric probands, suggesting that the pediatric spectrum of desminopathy reflects a distinct genetic architecture compared with adult-onset disease. These findings have direct implications for early diagnosis, risk stratification, and surveillance strategies in affected and at-risk children.

### Clinical and Genetic Features of Pediatric *DES* Cardiomyopathy

The present study offers new insights into the clinical and genetic characteristics of pediatric patients with *DES* variants. Pediatric *DES*-associated cardiomyopathy is exceedingly rare, with only 3 out of 413 children and adolescents with cardiomyopathies harboring P/LP *DES* variants in a recent European registry from 14 countries,^11^ and such cases are absent in several other large pediatric cardiomyopathy registries.^15–18^ In our study, pediatric patients accounted for 22.2% of all *DES* cases and were more frequently the proband within their families compared to adults, underscoring the early and often prominent clinical expression of *DES*-associated disease in childhood.

While cardiac involvement was the predominant feature in over half of the pediatric cases, approximately one in four presented with skeletal muscle symptoms, and a smaller subset had overlapping cardiac and musculoskeletal manifestations at first evaluation. This reinforces the importance of a multidisciplinary diagnostic approach, particularly in children presenting with muscle weakness, elevated creatine kinase or unexplained cardiac conduction abnormalities. The broad phenotypic spectrum observed in this cohort, including cases of dilated, hypertrophic, restrictive, and arrhythmogenic cardiomyopathies, mirrors the heterogeneity seen in adult *DES* cohorts and emphasizes the need for careful longitudinal phenotyping.^4^ Notably, even though our analysis required all included patients to have an imaging test performed, completion of cardiac magnetic resonance imaging in this population is often challenging due to the need for either sedation or general anesthesia. Thus, it is possible that the cardiomyopathy penetrance in this population is underestimated with ECG and echocardiogram alone.

While sustained VA and AV block are the abnormalities commonly surveilled for when managing patients with *DES*-associated cardiomyopathy, it is important to note that sinus bradycardia during the early stages of sinus node dysfunction and early-onset AF can be more subtle early presentations of the disease in the pediatric population, sometimes occurring before detectable cardiomyopathy. AF is exceedingly rare in the general pediatric population and always warrants a thorough evaluation. A high index of suspicion must be maintained in pediatric patients presenting with these more subtle manifestations, even in the absence of the neuromuscular abnormalities. This highlights the importance of genetic testing in pediatric patients with conduction system abnormalities and uncommon arrhythmias for age.

### Genotype Spectrum and Inheritance Patterns

The genetic landscape of pediatric cardiomyopathies remains inadequately studied, partially explained by their rarity and heterogeneity, with recent studies identifying causal variants in 40-50% of cases.^15,17–19^ Genetic evaluation is crucial for accurate diagnosis, prognostic assessment, and family counseling and holds promise for personalized management.^16,20^ Desminopathies typically follow an autosomal dominant inheritance pattern, but a small subset has an autosomal recessive transmission, and a significant number of variants arise *de novo*.^3,21^ We observed substantial heterogeneity in inheritance patterns and variant classes.

A notable proportion of patients, particularly probands, harbored biallelic (homozygous or compound heterozygous) or *de novo* variants, supporting a genotype– and inheritance–phenotype correlation in which highly disruptive allele states are associated with earlier onset and more severe disease. Although our cohort size limits statistical power for meaningful subgroup comparisons, these insights suggest the need for further investigation.

### Clinical Event Burden and Group Differences

Although adult and pediatric cardiomyopathies often have similar morphological and clinical manifestations, pediatric cardiomyopathies have worse outcomes,^9,10,22^ with nearly 40% of affected children either undergoing a heart transplantation or dying within two years of diagnosis.^23^ Clinical outcomes were poor in our pediatric *DES* patients: nearly 40% of patients developed CCD requiring CIED placement, one in five experienced sustained VA, and more than one-third suffered HF events during follow-up. Notably, CCD and HF events were more common in probands compared to affected pediatric family members, suggesting that earlier or more penetrant disease may be influenced by variant severity, inheritance pattern, or ascertainment bias. In contrast, the risk of sustained VA appeared comparable and yet substantial between groups, highlighting the unpredictable nature of arrhythmic complications in *DES* cardiomyopathy. Given that one in five pediatric *DES* patients died during follow-up, at a median age of 23.5 years, our data underscore the need for early recognition, timely intervention, and long-term risk management in affected children.

### Clinical Implications for Surveillance and Risk Management

Our findings raise important considerations for clinical care. First, surveillance of at-risk children should begin in early childhood and continue into adulthood, with periodic cardiac imaging, rhythm monitoring, and neurologic evaluation. Given the age-dependent penetrance observed in our cohort, vigilance during adolescence and young adulthood is particularly warranted. Second, although specific guidance for primary prevention ICD implantation in *DES* patients remains lacking, our results suggest that a substantial proportion of pediatric patients – both probands and family members – develop MACE, including life-threatening VA, within a few years of diagnosis. This supports the need for individualized risk assessment, and in cases of CCD, consideration of ICD over pacemaker implantation. When an ICD is considered, the high prevalence of CCD supports the use of a transvenous ICD with back-up pacing capabilities in this patient population. In contrast, subcutaneous or extravascular ICDs should only be considered in selected patients without CCD, given the strong preference to avoid transvenous leads and ensuing long-term risk for complications in the pediatric population, however, continued cardiac surveillance to screen for the development of CCD is mandatory in these patients. Third, the observed burden of *de novo* variants in over one-fifth of all pediatric probands, highlights the importance of *DES* genetic testing even in the absence of family history, especially when cardiac and neuromuscular features co-occur. It also highlights the critical need for cascade screening in families with a member diagnosed with *DES-*associated disease for early diagnosis in relatives and for establishing disease inheritance pattern, particularly in light of the relatively high rates of *de novo* and biallelic variants.

### Remaining Challenges and Future Directions

Despite these insights, several challenges remain. The phenotypic overlap with other neuromuscular or cardiomyopathic syndromes may delay diagnosis, particularly in cases with subtle early features. Clinical suspicion for *DES-*associated disease should be heightened in children presenting with cardiac and neuromuscular symptoms including muscle weakness and moderate CK elevation, cardiac conduction abnormalities, and cardiomyopathy, including dilated, restrictive, or less commonly hypertrophic or arrhythmogenic.^4^ Still, the diagnosis remains difficult due to the rarity of disease and variability in initial presentation. Additionally, there are no consensus guidelines for the management of pediatric *DES* cardiomyopathy, and decisions regarding surveillance intervals, device choice, or treatment initiation must currently be made on a case-by-case basis after shared decision making. Larger registries and prospective studies are needed to define gene- and variant-specific predictors of disease progression and distinct clinical outcomes.

### Study Strengths and Limitations

To our knowledge, this is the first study to date on pediatric patients with pathogenic or likely pathogenic *DES* variants. However, the systematic review design and variability in reporting across included studies limit the overall evidence quality. Reliance on published data restricted detailed characterization of skeletal myopathy and cardiomyopathy phenotypes and precluded analysis of medication effects, though no therapies have yet been shown to alter outcomes in *DES*-associated cardiomyopathy. There is also the potential for publication bias toward more severe phenotypes since pediatric patients with mild *DES* forms may not be published. However, due to the inclusion criteria in the present study, there may also be underreporting of patients with *DES-*associated disease who experienced SCD and were diagnosed post-mortem or in whom the diagnosis was never reached.

Since only living patients were included, the reported rates of ventricular arrhythmias and mortality may underestimate true event burden. Phenotypic differences between probands and relatives could reflect publication and ascertainment biases. Finally, our findings were derived from clinically diagnosed cases and should not be directly extrapolated to individuals with secondary findings or incidentally identified *DES* variants as their penetrance and prognosis remain unclear.^24,25^

## Conclusions

Pediatric patients with *DES*-associated cardiomyopathy exhibit early-onset, multisystem disease with heterogeneous cardiac phenotypes and a high burden of adverse outcomes. Cardiomyopathy penetrance begins as early as prepubertal age and increases through adolescence and early adulthood, underscoring the critical need for early initiation and longitudinal cardiac evaluation in at-risk children. Probands are more likely to harbor *de novo* or biallelic variants and experience conduction disease and HF events, though arrhythmic risk appears to be similar and substantial in pediatric relatives. These findings highlight the importance of early genetic testing in children presenting with unexplained cardiomyopathy or neuromuscular features, even in the absence of family history. Genotype-informed risk assessment and tailored multidisciplinary care is critical for improving outcomes in this high-risk population. Further studies are needed to identify gene-specific predictors of distinct outcomes and develop genotype-informed management protocols.

## Sources of Funding

The Johns Hopkins ARVC Program is supported by the Leonie-Wild Foundation, the Leyla Erkan Family Fund for ARVD Research, The Hugh Calkins, Marvin H. Weiner, and Jacqueline J. Bernstein Cardiac Arrhythmia Center, the Dr. Francis P. Chiramonte Private Foundation, the Dr. Satish, Rupal, and Robin Shah ARVD Fund at Johns Hopkins, the Bogle Foundation, the Campanella family, the Patrick J. Harrison Family, the Peter French Memorial Foundation, and the Wilmerding Endowments. Dr. Asatryan was supported by the 2022 Research Fellowship for aspiring electrophysiologists from the Swiss Heart Rhythm Foundation, and a postdoctoral research fellowship grant from the Gottfried und Julia Bangerter-Rhyner-Stiftung (Switzerland). Dr. Rieder was funded by a grant from the Gottfried und Julia Bangerter-Rhyner-Stiftung (Switzerland) and a grant from the Inselspital Bern (Nachwuchsförderungs-Grant). Dr Carrick was funded by an NIH T32 Grant (T32HL007227), the NIH LRP (L30HL165535), and is a recipient of the Semyon and Janna Friedman Family Fellowship Award. Dr. Te Riele is supported by a grant from ZonMW (2024 Clinical Fellows grant no. 09032232310042) and a HORIZON Cardiogenomics pathfinder (IMPACT, grant no. 101115536).

## Disclosures

BA received support from Abbott and Boston Scientific for attending meetings and travel unrelated to this work. AG received advisory board / consulting fees from Lexeo and has served as an unpaid consultant for StrideBio. AtR is a consultant for Tenaya therapeutics, Rocket Pharmaceutical and Lexeo for unrelated work. HC is a consultant for Medtronic Inc., Biosense Webster, Pfizer, Rocket, and Abbott. CAJ receives research funding from Lexeo Therapeutics, Rocket Pharmaceuticals, Tenaya Therapeutics, and ARVADA Therapeutics for unrelated work; SM and CT have received salary support from these companies. CAJ is a past consultant for Lexeo Therapeutics. All other authors have reported that they have no relationships relevant to the contents of this paper to disclose. The other authors report that they have no relevant conflicts of interest to disclose.

## Non-standard Abbreviations and Acronyms

AF: atrial fibrillation
ARVC: arrhythmogenic right ventricular cardiomyopathy
CCD: cardiac conduction disease
DCM: dilated cardiomyopathy
HCM: hypertrophic cardiomyopathy
HF: heart failure
MACE: major adverse cardiac events
RCM: restrictive cardiomyopathy
SCD: sudden cardiac death
VA: sustained ventricular arrhythmia

## Data Availability

The dataset generated through the extraction of data from published studies will be made available upon reasonable request.

## Notes

### Author Declarations

The data used in the systematic review and individual patient data meta-analysis are publicly available and ethics approval was not required.

